# The King’s College London Coronavirus Health and Experiences of Colleagues at King’s Study: SARS-CoV-2 antibody response in an occupational sample

**DOI:** 10.1101/2020.09.10.20191841

**Authors:** Daniel Leightley, Valentina Vitiello, Gabriella Bergin-Cartwright, Alice Wickersham, Katrina A.S. Davis, Sharon A.M Stevelink, Matthew Hotopf, Reza Razavi, On behalf of the KCL CHECK research team

## Abstract

We report test results for SARS-CoV-2 antibodies in an occupational group of postgraduate research students and current members of staff at King’s College London. Between June and July 2020, antibody testing kits were sent to n=2296 participants; n=2004 (86.3%) responded, of whom n=1882 (93.9%) returned valid test results. Of those that returned valid results, n=124 (6.6%) tested positive for SARS-CoV-2 antibodies, with initial comparisons showing variation by age group and clinical exposure.

## Main

Establishing prevalence of SARS-CoV-2 poses significant challenges to the research community in light of social distancing measures which can preclude in-person testing across population cohorts. Antibody tests can be used to determine whether a person has already had SARS-CoV-2. Specific IgM and or IgG antibodies should be detectable from 4-5 days post-infection, with positive IgM antibodies in 70% of symptomatic patients by day 8-14 and 90% of antibody tests positive by day 11-24 [1]. We investigated the feasibility of remote home antibody testing as part of large-scale study monitoring the effects of SARS-CoV-2. A detailed analysis of home testing with Rapid Immunoglobulin Test Cassettes is reported in [2].

The King’s College London Coronavirus Health and Experiences of Colleagues at King’s Study (KCL CHECK) seeks to explore the psychological, social and physical impact of SARS-CoV-2 in a longitudinal cohort of postgraduate research students and current members of staff at King’s College London, a large Russell Group University in London, UK (for study protocol see [3]). A total of n=2416 participants are enrolled in the KCL CHECK Study, with n=2296 (95.0%) providing consent for SARS-CoV-2 antibody home testing. Tests were posted to participants in June and July 2020. More than 70% of participants who took part in testing were based in the South of England (Figure 1). Approximately n=224 (9.8%) participants needed their home testing kits to be resent due to technical issues with the test, an invalid result or loss in transit.

**Figure 1.**
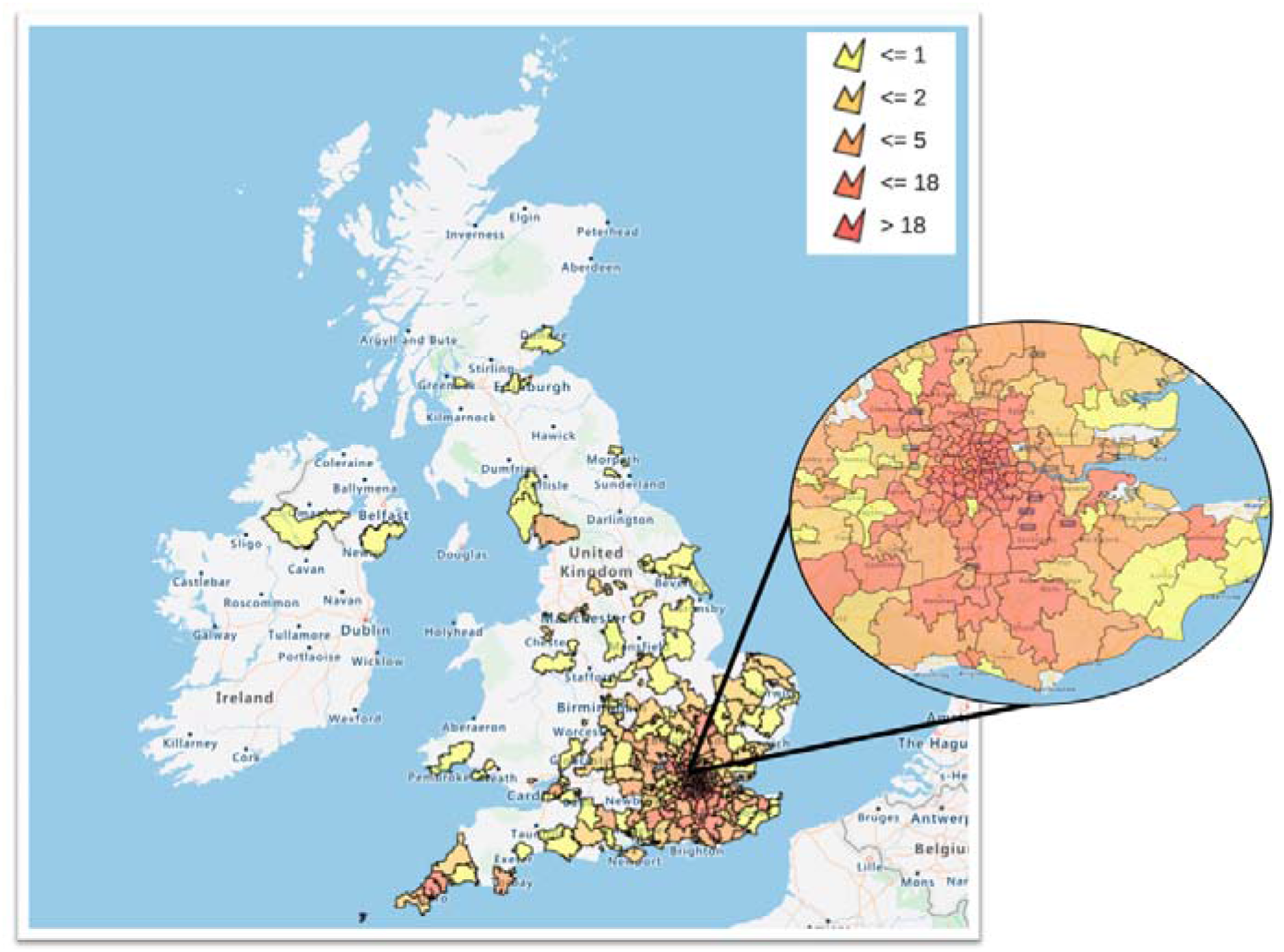
Heatmap indicates participants who took part in testing. Participants were aggregated into parliamentary constituency based on the first 3 digits of their postcode.

The characteristics of participants who provided a valid SARS-CoV-2 antibody results are summarised in Table 1. Nonetheless, n=2004 (86.3%) participants successfully returned a test result via photograph, of which n=1882 (93.9%) were valid tests. Overall, n=1758 (93.4%) were negative and n=124 (6.6%) were positive. Stratifying by occupational characteristics showed some variation in positive rates among staff (n=90; 6.1%), students (n=25; 8.0%) and those fulfilling both a student and staff role (n=9; 11.7%). Similarly, there was some variation in positive rates based on not having a clinical role (n=111; 6.5%), having a clinical role without suspected Covid-19 patient contact (n=7; 7.1%) and having a clinical role with suspected Covid-19 patient contact (n=5; 9.1%).Figure 1

**Table 1.**
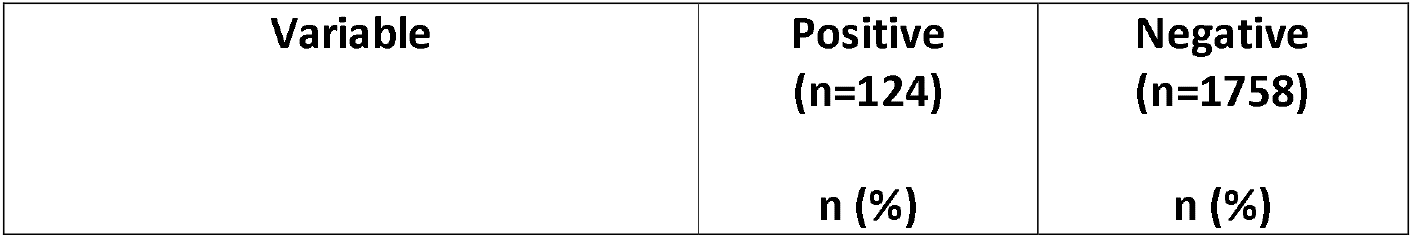
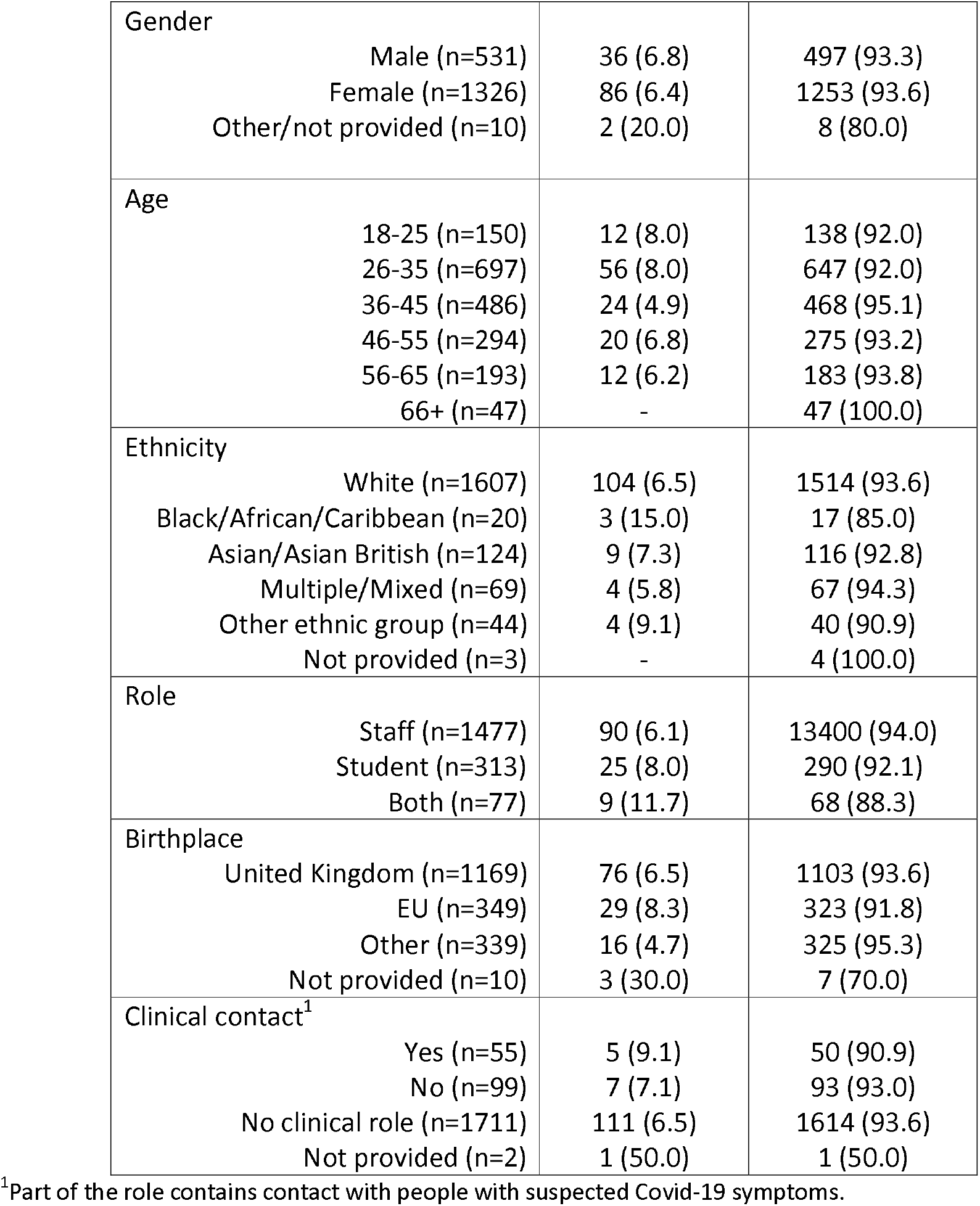
SARS-CoV-2 antibody results stratified by self-reported socio-demographic information. N=1882.

| Variable | Positive<br>(n=124) | Negative<br>(n=1758) |
| --- | --- | --- |
|  | n (%) | n (%) |

|  |  |  |  |
| --- | --- | --- | --- |
| Gender | Male (n=531) | 36 (6.8) | 497 (93.3) |
|  | Female (n=1326) | 86 (6.4) | 1253 (93.6) |
|  | Other/not provided (n=10) | 2 (20.0) | 8 (80.0) |
| Age | 18-25 (n=150) | 12 (8.0) | 138 (92.0) |
|  | 26-35 (n=697) | 56 (8.0) | 647 (92.0) |
|  | 36-45 (n=486) | 24 (4.9) | 468 (95.1) |
|  | 46-55 (n=294) | 20 (6.8) | 275 (93.2) |
|  | 56-65 (n=193) | 12 (6.2) | 183 (93.8) |
|  | 66+ (n=47) | - | 47 (100.0) |
| Ethnicity | White (n=1607) | 104 (6.5) | 1514 (93.6) |
|  | Black/African/Caribbean (n=20) | 3 (15.0) | 17 (85.0) |
|  | Asian/Asian British (n=124) | 9 (7.3) | 116 (92.8) |
|  | Multiple/Mixed (n=69) | 4 (5.8) | 67 (94.3) |
|  | Other ethnic group (n=44) | 4 (9.1) | 40 (90.9) |
|  | Not provided (n=3) | - | 4 (100.0) |
| Role | Staff (n=1477) | 90 (6.1) | 13400 (94.0) |
|  | Student (n=313) | 25 (8.0) | 290 (92.1) |
|  | Both (n=77) | 9 (11.7) | 68 (88.3) |
| Birthplace | United Kingdom (n=1169) | 76 (6.5) | 1103 (93.6) |
|  | EU (n=349) | 29 (8.3) | 323 (91.8) |
|  | Other (n=339) | 16 (4.7) | 325 (95.3) |
|  | Not provided (n=10) | 3 (30.0) | 7 (70.0) |
| Clinical contact<sup>1</sup> | Yes (n=55) | 5 (9.1) | 50 (90.9) |
|  | No (n=99) | 7 (7.1) | 93 (93.0) |
|  | No clinical role (n=1711) | 111 (6.5) | 1614 (93.6) |
|  | Not provided (n=2) | 1 (50.0) | 1 (50.0) |

<sup>1</sup>Part of the role contains contact with people with suspected Covid-19 symptoms.

Our study has some limitations. The SureScreen Diagnostics COVID-19 IgG/IgM Rapid Test Cassette used in this study was designed for “point-of-care” testing and, at the time of testing, rapid test cassettes had been certified by the Medicines and Healthcare products Regulatory Agency for use in laboratories using venous blood as there was insufficient data on their reliability when using capillary blood. The test which was performed by the study participants was therefore “off label”, which may explain the number of invalid results. Second, we relied on participants providing a clear photograph of the cassette result and accurate test times to ensure the picture was taken within the timeframe where the result is known to be reliable. Finally, the duration which antibodies remain in the body is unknown, and there is uncertainty about the utility of antibody testing for seroprevalence surveys for public health management purposes [4]. We plan to repeat the antibody testing at regular intervals, allowing further evaluation of antibody persistence over time.

Nonetheless, our initial findings indicate a similar positive rate in our cohort compared to the UK general population [5]. However, this is lower than a recent report which found the South and London had a prevalence of 13%, perhaps surprising considering the proportion of staff in our cohort working in a clinical setting (9.9%) [5]. With 93.9% of participants returning a valid photograph, our study highlights the feasibility of rapidly deploying low-cost SARS-CoV-2 serological testing without the need for face-to-face contact in an occupational setting.

## Methods

### Study design

The KCL CHECK Study is a longitudinal cohort study of the health and wellbeing of postgraduate research students and current members of staff at King’s College London. Participants were recruited by email and volunteered to complete surveys and antibody testing over the 18-month study period. While there was no incentivisation, participation could have been motivated by the offer of testing. A Rapid Immunoglobulin Test Cassette was used to detect the presence of IgM and IgG antibodies to the ‘spike’ protein, thereby providing evidence for previous infection with SARS-CoV-2. SureScreen Diagnostics COVID-19 IgG/IgM Rapid Test Cassettes, the necessary equipment and detailed instructions were sent to participants’ home address. Participants were informed that the test was not licenced for clinical use and was being used for research purposes. A reporting sheet included each participant’s unique identifier, such that when participants submitted the photograph online via the study website, it could be securely linked to their survey results. Participant photographs were then analysed by the research team and rated according to the following scales: ‘positive’ – indicating the presence of IgG/IgM (denoted via pink lines on each item), negative (denoted via a pink line on the control item) and invalid (denoted by no lines appearing on any item or blood in the buffer zone). Inter-rater reliability (Cohen’s kappa) was κ=0.84 (95% CI: 0.87-0.97), agreement 95.18% indicating strong agreement [6].

### Statistical analysis

Statistical analysis was performed in STATA 16.0. Descriptive statistics and antibody test results are reported as frequency and unweighted row percentages.

## Data Availability

Researchers may apply to have access to pseudonymised data. Requests to access study data is subject to submission of a research proposal to the Principal Investigators (Professor Matthew Hotopf, Professor Reza Razavi and Dr Sharon Stevelink). All requests must be made in accordance with the UK Policy Framework for Health and Social Care research. Where the applicant is outside of Kings College London, a data-sharing agreement is required. Raw data presented in this study is provided in the Supplementary Dataset.

## Acknowledgements

The research team would like to thank Jonathan Edgeworth for support during study setup. The research team would like to acknowledge Liam Jones, Lisa Sanderson, Jana Kim and Laila Danesh for providing support during packaging of the home testing kits. We would also like to thank Charlotte Williamson for providing support in annotating test result photographs. This paper represents independent research part funded by the National Institute for Health Research (NIHR) Biomedical Research Centre at South London and Maudsley NHS Foundation Trust and King’s College London. The views expressed are those of the author(s) and not necessarily those of the NHS, the NIHR or the Department of Health and Social Care.

## Ethical approval

Ethical approval has been gained from King’s College London Psychiatry, Nursing and Midwifery Research Ethics Committee (HR-19/20-18247). Participants provided informed consent to take part.

## Data availability

Researchers may apply to have access to pseudonymised data. Requests to access study data is subject to submission of a research proposal to the Principal Investigators (Professor Matthew Hotopf, Professor Reza Razavi and Dr Sharon Stevelink). All requests must be made in accordance with the UK Policy Framework for Health and Social Care research. Where the applicant is outside of King’s College London, a data-sharing agreement is required. Raw data presented in this study is provided in the Supplementary Dataset.

## References

[1] N. J. Beeching, T. E. Fletcher, and M. B. J. Beadsworth, “Covid-19: testing times,” BMJ, p. m1403, Apr. 2020, doi: 10.1136/bmj.m1403.

[2] S. Pickering et al., “Comparative assessment of multiple COVID-19 serological technologies supports continued evaluation of point-of-care lateral flow assays in hospital and community healthcare settings,” PLOS Pathog., vol. 16, no. 9, p. e1008817, Sep. 2020, doi: 10.1371/journal.ppat.1008817.

[3] K. A. S. Davis et al., “The King’s College London Coronavirus Health and Experiences of Colleagues at King’s Study (KCL CHECK) protocol paper: a platform for study of the effects of coronavirus pandemic on staff and postgraduate students.,” medRxiv, p. 2020.06.16.20132456, Jan. 2020, doi: 10.1101/2020.06.16.20132456.

[4] J. J. Deeks et al., “Antibody tests for identification of current and past infection with SARS-CoV-2,” Cochrane Database Syst. Rev., Jun. 2020, doi: 10.1002/14651858.CD013652.

[5] H. Ward et al., “Antibody prevalence for SARS-CoV-2 following the peak of the pandemic in England: REACT2 study in 100,000 adults,” medRxiv, 2020.

[6] M. L. McHugh, “Interrater reliability: the kappa statistic.,” Biochem. medica, vol. 22, no. 3, pp. 276–82, 2012, [Online]. Available: http://www.ncbi.nlm.nih.gov/pubmed/23092060.

